# Monitors to Improve Indoor Carbon Dioxide Concentrations in the Hospital: Background, Rationale and Protocol for a Randomized, Sham-controlled, Cross-Over, Open Label Trial

**DOI:** 10.1101/2021.05.08.21256868

**Authors:** Michaël R. Laurent, Johan Frans

**Affiliations:** Geriatrics Department, Imelda Hospital, Bonheiden, Belgium; Department of Medical Microbiology, Imelda Hospital, Bonheiden, Belgium

**Author notes:** **Correspondence:** Dr. Michaël R. Laurent, MD PhD, Imeldalaan 9, 2820 Bonheiden, Belgium. **Author contribution statement:** Conceptualization (equal), Resources (equal), Data curation (ML), Formal analysis (ML), Investigation (equal), Methodology (equal), Project administration (ML), Writing - original draft (equal), Writing - review & editing (equal), Approval of the final manuscript (equal). **Ethics approval statement:** The study protocol was reviewed and approved by the Institutional Review Board of Imelda Hospital, Bonheiden, Belgium. **Patient consent statement:** not applicable. **Data availability statement:** Following publication, all data supporting this manuscript will be made available to established investigators upon simple request. **Funding statement:** The authors report no external funding related to this work. **Permission to reproduce material from other sources:** not applicable.

**Keywords:** Carbon dioxide, coronavirus disease 2019, Geriatrics, Healthcare-associated Infections, Hospitals, Ventilation

## Abstract

Coronavirus disease 2019 (COVID-19) has caused considerably morbidity and mortality worldwide, mainly among older adults. Hospital outbreaks contribute to the burden of this disease, despite optimal hand hygiene and personal protective equipment such as masks and face shields. Ventilation with fresh outdoor air has emerged as an important strategy to reduce indoor aerosol transmission of COVID-19. Carbon dioxide (CO_2_) monitors are increasingly advocated to facilitate ventilation in schools, long-term care facilities, offices and public buildings. Moreover, several health authorities have issued guidelines for target CO_2_ values in work as well as clinical environments. Given that modern hospitals have superior indoor air quality control systems, it remains however unknown whether feedback from CO_2_ monitors is needed and/or effective to improve ventilation further. Here, we describe the rationale and protocol for a randomized, sham-controlled, crossover, open label trial of CO_2_ monitors in double-bed hospital rooms in two acute geriatric wards. Based on pilot data, Aranet4 Home^®^ monitors will be used to alert nurses and other staff to raised indoor CO_2_ concentrations. Practical limitations in implementing CO_2_ monitors are discussed, and will be surveyed among staff as additional study outcomes. The Monitors to Improve Indoor Carbon Dioxide (CO_2_) Concentrations in the Hospital (MICH) trial is registered at ClinicalTrials.gov, identifier: <u>NCT04770597</u>.

## 1. INTRODUCTION

The coronavirus disease 2019 (COVID-19) pandemic has posed a high burden on societies, healthcare systems and long-term care facilities worldwide. COVID-19 has caused considerably morbidity and mortality, particularly in frail older adults [1], [2]. While there is little direct evidence for any transmission route, there is accumulating evidence for respiratory transmission of severe acute respiratory syndrome coronavirus 2 (SARS-CoV-2, the virus which causes COVID-19), not only via droplets but also via smaller particles in aerosols, mainly in closed spaces [3], [4], [5].

Nosocomial transmission contributes to the incidence and mortality of COVID-19 [2], [6], [7]. Acute medical and surgical wards in particular host many older adults close together. Therefore, unprecedented infection control measures have been implemented in hospitals since 2020. These include universal mask wearing by patients and caregivers, increased attention to hand and surface disinfection, and screening patients and/or staff for SARS-CoV-2 [8]. Other measures include physical distancing among staff and use of mobile high-efficiency particulate air (HEPA)-filters. Nevertheless, clusters of SARS-CoV-2 transmission have been documented despite all these countermeasures [6]. Exposure to an infected patient for at least 15 minutes is a documented risk factor in this regard [6].

Ventilation with fresh outdoor air is increasingly highlighted as a strategy to avoid indoor aerosol transmission of respiratory pathogens [9]. One convenient surrogate parameter for ventilation in this context, is the indoor carbon dioxide (CO_2_) concentration [10]. Humans exhale CO_2_ at concentrations of almost 40 000 p.p.m. [11], compared to outdoor concentrations which have increased in recent years to values averaging ∼450 p.p.m. (fluctuating between 400-500 p.p.m.). Thus, in the absence of combustion, animals and certain chemical reactions (which also produce CO_2_) or plants (which catalyze CO_2_ during photosynthesis), the increase in CO_2_ concentrations above outdoor levels, reflects the indoor concentration of human exhaled air (potentially carrying respiratory pathogens). The total amount of CO_2_ which humans exhale is determined by their respiratory minute volume or oxygen consumption (which correlates with physical activity) and their metabolic respiratory quotient (ratio of CO_2_ exhaled to oxygen consumed, which is influenced by diet and metabolic factors). Rudnik & Milton reported that under average circumstances, an indoor CO_2_ concentration > 380 p.p.m. above outside levels implies that inhaled air contains 1% exhaled air [11]. In turn, this rebreathed fraction or “shared air” correlates with the basic reproductive number of respiratory infections [11]. On the other hand, increased ventilation (particularly displacement ventilation [12]) could also facilitate spreading of pathogens if the air flows from the contaminant source towards susceptible, unmasked individuals [13], [14].

There is no universal agreement on optimal cut-offs for indoor CO_2_ concentrations in general, or in the context of COVID-19 in particular. Regulatory bodies in several countries have proposed guidelines to maintain indoor CO_2_ concentrations below values typically ranging from 800 to 1000 p.p.m. [15], [16], [17]. One COVID-19 cluster in a Dutch nursing home was associated with the use of an energy-saving CO_2_-driven ventilation system, which maintained indoor CO_2_ concentrations around 1000 p.p.m. [18]. It should be noted that indoor air quality standards for buildings sometimes refer to *average* CO_2_ levels over *prolonged* time periods (*e*.*g*. 8 hours [19]), although the risk of respiratory infections also applies to shorter exposures [6], [11].

As the COVID-19 pandemic continues, CO_2_ monitors are increasingly deployed and recommended (for example, in countries like Germany or Norway [17]) to monitor ventilation and prevent aerosol transmission of SARS-CoV-2 in schools, long-term care facilities, offices and public buildings [15], [17], [20]. Some experts have called for the use of CO_2_ monitors by nurses in hospitals and nursing homes too [20]. However, empirical evidence supporting the use of CO_2_ monitors to improve ventilation (let alone to prevent COVID-19) in any setting is lagging behind.

Modern hospitals, particularly operating and delivery rooms, intensive care units and microbiology laboratories, are equipped with superior heating, ventilation and air conditioning (HVAC) systems, which also remove particles via HEPA filters. For normal patient rooms however, the recommended outdoor and total air change rates per hour are only 2 and 4, respectively [21]. While these rates may be sufficient to meet indoor air quality requirements for patients, they may be temporarily insufficient when patient rooms are crowded by visitors or healthcare staff. Indeed, physical distancing and limiting occupant density, which are commonly recommended in stores, offices and public buildings, are challenging in hospitals, since patients depend on caregivers. In one study, CO_2_ production in hospital rooms correlated with room entries (indicated by infrared beam breaks) [22]. Moreover, opening windows in hospital may be avoided due to thermal and draft discomfort [13], [23] or risk of patient injury from falling out the window [24].

Few studies have actually reported indoor CO_2_ concentrations in hospitals [13]. Inappropriately high CO_2_ concentrations have been reported in hospitals without modern HVAC systems in Brazil (reaching peaks over 3000 p.p.m) [25] and China (daily averages > 1000 p.p.m.) [23]. In contrast, excellent CO_2_ values were reported in an Iranian intensive care unit [26] and a new hospital building in the United States (daily *average* CO_2_ < 150 p.p.m. above outside levels) [22]. A French study reported maximal CO_2_ concentrations of 1121 to 1325 p.p.m. in a hospital nursing care room and plaster room, respectively [27]. In a Taiwanese four-bed intensive care unit room, CO_2_ levels (range 828-1570 p.p.m.) were above 1000 p.p.m. during visitor hours 92% of the year [28]. Another study from Taiwan reported that among hospital sites, patient wards had the highest average CO_2_ concentration (1063 ± 483 p.p.m., N=3 hospitals) [29]. Thus, it is clear that despite guidelines for hospital HVAC systems and CO_2_ targets in buildings, ventilation in regular wards may be worse than commonly appreciated. Most studies have reported daily average CO_2_ levels or maximal levels during short measurement periods. More research is needed not only to provide a more detailed overview of indoor CO_2_ fluctuations in hospital rooms, but even more so to define optimal strategies to maintain CO_2_ below recommended maximum levels. Interestingly, Yang *et al*. reported daily peaks > 1000 p.p.m. in the hospital environment, which could be mitigated using an integrated monitoring system which alerted medical supervisors and automatically activated ventilation [19].

Given this background, the authors launched a prospective interventional study to assess the efficacy and feasibility of monitors to maintain indoor CO_2_ concentrations in their hospital below 800 – 1000 p.p.m. Here, we describe the rationale and pilot data supporting the design and protocol of the “<u>M</u>onitors to Improve Indoor <u>C</u>arbon Dioxide <u>C</u>oncentrations in the <u>H</u>ospital” (MICH) randomized trial.

## 2. PROTOCOL

This trial protocol (version 2.0, April 9^th^, 2020) is reported in accordance with the Standard Protocol Items: Recommendations for Interventional Trials (SPIRIT) 2013 statement [30]. The SPIRIT checklist is provided (**Supplementary Table 1**).

### 2.1. Study design

This study is a randomized, sham-controlled, open-label, crossover superiority trial, performed consecutively in two acute geriatric wards. In each ward, six double bed rooms will be fitted with Aranet4 Home^®^ CO_2_ monitors, which constitute the unit of randomization. The crossover design involves three phases, each lasting seven days (**Figure 1**).

**Figure 1:**
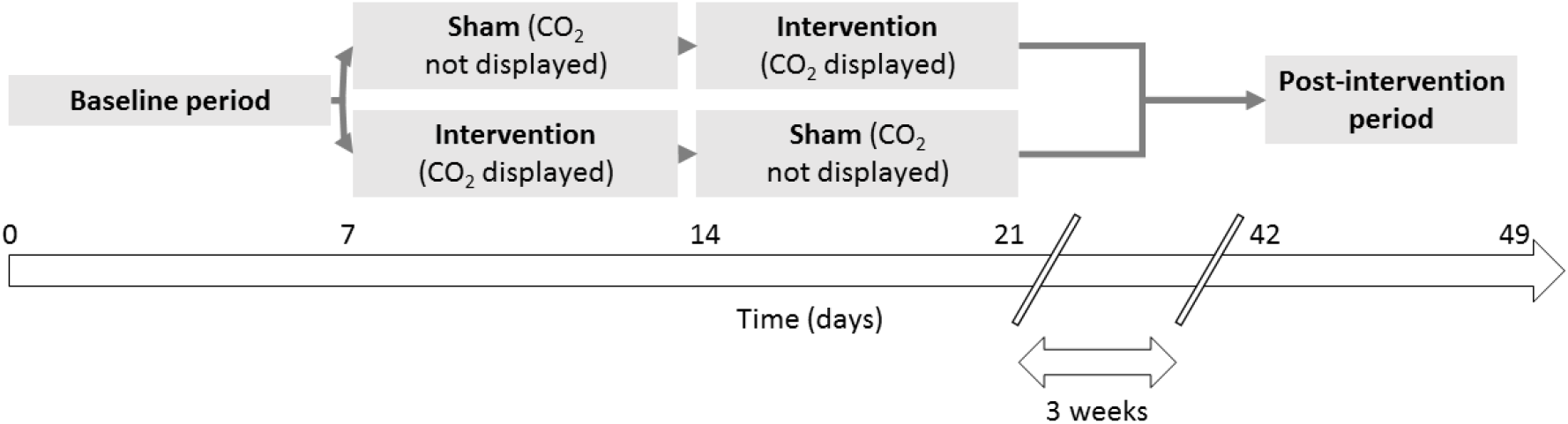
Study design overview. 1. **Pre-intervention (baseline) period (1 week):** all monitors facing downwards, with staff blinded to measurements. 2. **Intervention period (2 x 1 weeks):** Each monitor will be randomized either to be visible to the clinical staff for one week, or in the sham-controlled group, to be turned with the screen facing downwards for one week. After one week, the monitors cross over to the other group (AB/BA crossover design) for one week. After two weeks, the sensors will be removed from the first ward and moved for three weeks to the second ward. Thus, the duration of the trial is seven weeks per ward, and ten weeks in total. 3. **Post-intervention period (1 week):** after a three week interval, the sensors are again installed in six double-bed rooms, with their display facing downwards and staff blinded to the measurements.

Given the open-label design, the sham-control group was chosen to take potential observation bias (Hawthorne effect) and carryover effects into account. The primary and secondary endpoints will be compared between the active and sham-controlled groups in the intervention phase. However, because we expect some carry-over effects from the intervention to the sham groups, pre-planned analyses will compare the intervention against the pre-intervention phase. Furthermore, comparison between the intervention and post-intervention phase will allow us to investigate whether significant wear-off effects can be demonstrated or not.

Sensors will be randomized by the investigators to the sham/intervention or intervention/sham arm respectively, immediately prior to the intervention using an online random sequence generator (1:1 ratio, N = 1 block size). Due to the nature of the intervention, allocation concealment is not possible. Investigators performing the analysis will also not be blinded to intervention groups. However, the *a priori* statistical analysis plan and the availability of data for independent review by other scientists mitigates the risk of bias from this source. Data will be analyzed using the intention-to-treat principle, *e*.*g*. even when the monitors in the sham group are unblinded or *vice versa*.

### 2.2. Setting

This single-center trial will run at the Geriatrics Department, which is located in the oldest wing of Imelda general hospital in the rural area of Bonheiden, Belgium. The hospital is surrounded by nature and there is no traffic or other nearby CO_2_ source. Our department has 69 acute beds (single- and double-bed rooms) divided over three wards, and one rehabilitation ward with 20 beds (transformed into a dedicated COVID-19 unit during the study). Among general acute wards with standard air exchange rates, the Geriatric Department was selected for several reasons. First, bed occupancy rate in the department is typically > 90 %. Secondly, almost all patients require assistance from staff for activities of daily living such as washing, dressing, meals *etc*. Thirdly, geriatrics has higher staffing levels than other acute wards and many nursing students (up to 10/ward). All these elements increase room crowding.

On the other hand, many older patients have low levels of physical activity and thus low respiratory minute volumes. Visitors were also allowed very restrictively due to the ongoing COVID-19 epidemic, typically only 1 hour/week during most of the study period (with exceptions for patients receiving palliative care, which are however typically housed in single rooms). The latter elements may be associated with lower indoor CO_2_ concentrations.

For this study, we selected two acute wards with near-identical architecture, on the first and second floor (our third ward having slightly more spacious rooms). All rooms recently had air conditioning installed (cooling mode only). Outdoor CO_2_ concentrations will be measured on an unused terrace immediately adjacent to the wards. The position of the sensors, along with the location and size of the respective double-bed rooms is illustrated in **Figure 2**. The patient rooms all had a similar surface area of ∼20-21 m² and a 3-4 m² bathroom. The doctor’s office is 23.5 m² and hosts 2 geriatricians, 1-2 registrars and 2-4 trainees.

**Figure 2:**
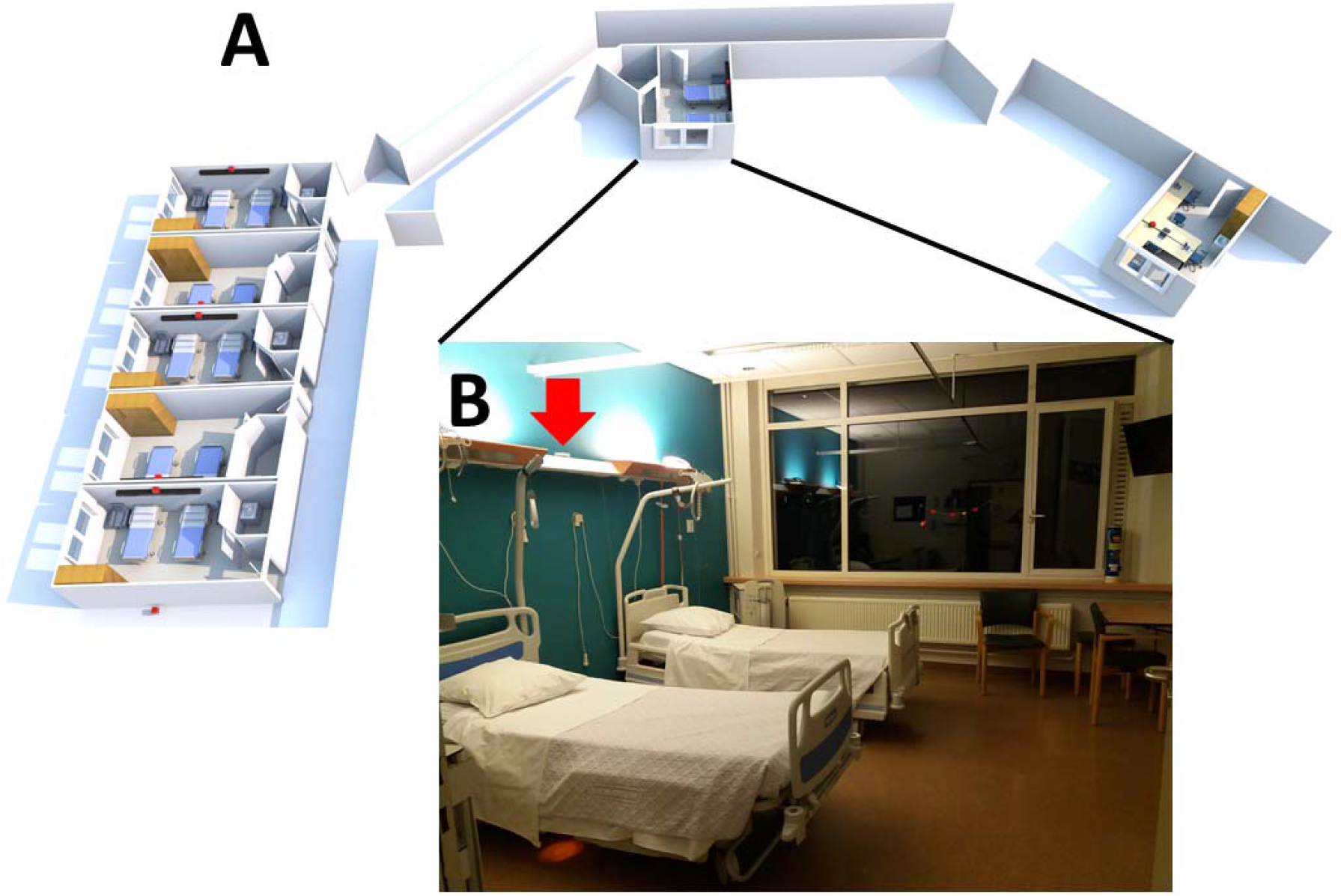
Position of the sensors and double-bed rooms during the study. **A:** Three-dimensional representation of the ward showing the position of the six sensors (small red squares) in the double-bed rooms, the doctor’s office (*right*) and the outdoor CO_2_ sensor (*bottom left*). **B (inset):** Representative room photograph. The arrow indicates the position of the sensor on the overhead light box.

### 2.3. Intervention

This study will use six Aranet4 Home^®^ and one Airthings Wave Plus^®^ monitor (measurement ranges 0 – 9999 p.p.m. and 400 – 5000 p.p.m., respectively). Both monitors are commercially available in Europe and use non-dispersive infra-red CO_2_ sensors with ± 3 % accuracy reported by the manufacturer, and raw data logging with time stamps. The Aranet^®^ sensors are factory-calibrated, recommended for use in schools by the Federation of European HVAC Associations [31], and have been shown to be reliable compared to research-grade instruments and to reflect ventilation rates in a clinical environment [32]. The Airthings^®^ instrument was used after the seven-day self-calibration period. When we cross-calibrated our devices (*i*.*e*., putting all sensors next to each other), they displayed near-identical values. We also compared the CO_2_ values from Aranet4 Home^®^ to a professional indoor climate multimeter (Testo^®^ 435-1) which showed identical measurement values. The sensors will be placed in the room at a height between 1 and 2 meters and not near the window or door.

Pilot data suggested that the highest CO_2_ concentrations were observed in patient rooms (**Figure 3**, and other data not shown), in line with previous literature [27]. When measured in the doctor’s office, CO_2_ values peaked during office hours but remained < 1000 p.p.m. (median 601 p.p.m., range 453 – 984 p.p.m.). We selected the Aranet4 Home^®^ for use in the patient rooms because, in contrast to the Airthings Wave Plus^®^, it has a display showing current CO_2_ levels and because it can be set to bleep when threshold values are exceeded. Moreover, it proved to be more responsive, which was partly but not completely explained by its superior time-resolution (measurements possible at one-minute intervals, compared to 5 minutes with Airthings Wave Plus^®^), as shown in **Figure 3B**. Median (range) CO_2_ values were 653 p.p.m. (541 – 1367 p.p.m.) with the Airthings Wave Plus^®^ *vs*. 689 p.p.m. (548 – 1540 p.p.m.) with Aranet4 Home^®^, difference in medians -36 p.p.m. (95% confidence interval -41 to -32, P < 0.0001 by Wilcoxon matched-pairs signed rank test). The values were not normally distributed (P < 0.0001 D’Agostino-Pearson omnibus K2 test) and significantly paired (Spearman rs 0.9344, P < 0.0001). The Aranet^®^ monitor data will be analyzed using two-minute bins due to ongoing technical problems with the stability of the Bluetooth^®^ connection to Android^®^ devices. We will use the Airthings Wave Plus^®^ to monitor outside CO_2_ levels continuously during the trial.

**Figure 3.**
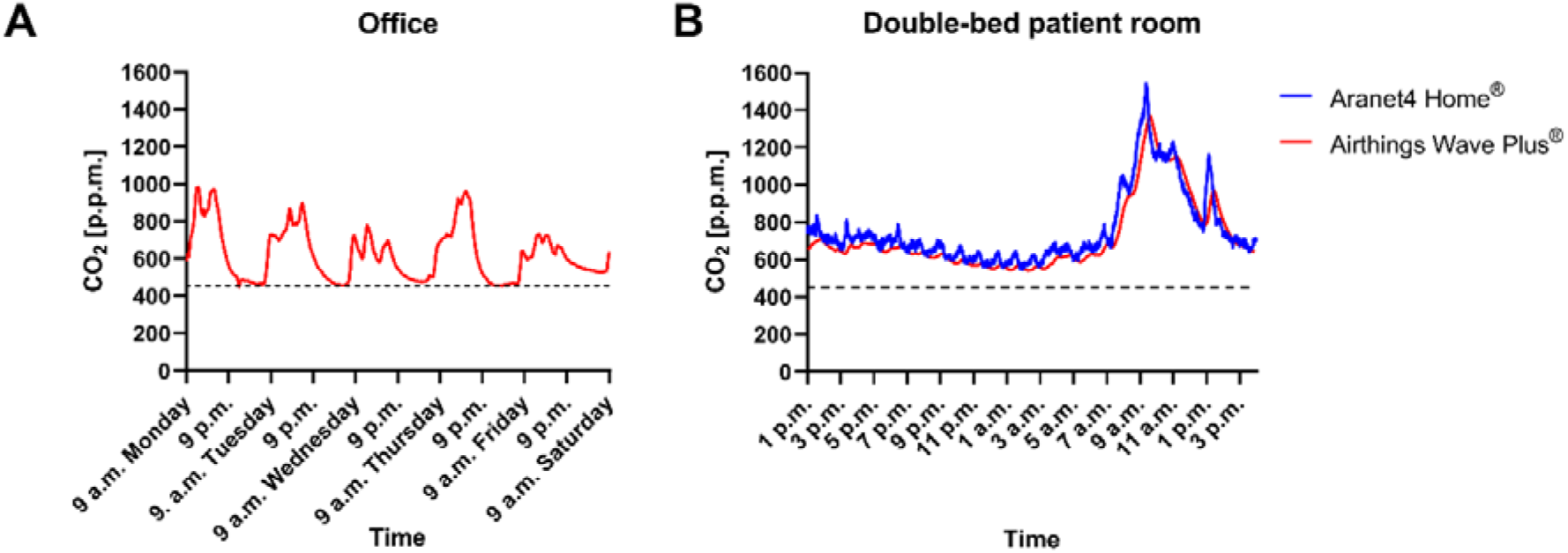
**A:** Indoor CO_2_ measured using Airthings Wave Plus^®^ sensor in the doctor’s office (non-patient area) during one working week. CO_2_ levels increased during office hours and returned to outside CO_2_ levels (*dashed line*) by next morning. One window and door remained almost constantly open. Staff were blinded and unaware of measurements. **B:** Pilot data showing indoor CO_2_ concentrations in a double-bed patient room over a 27-hour period, measured simultaneously with an Aranet4 Home^®^ and an Airthings Wave Plus^®^ monitor. A slight delay can be seen as an offset with the Airthings Wave Plus^®^ sensor. Staff were blinded to measurements. Dashed line indicates median outdoor CO_2_ concentrations.

At the start of the randomized intervention period, clinical staff on each ward will be educated by the principal investigator on the purpose and methods of the trial and strategies to improve ventilation. During the two-week intervention period, staff will be interviewed at least every three days regarding difficulties in achieving CO_2_ targets, and additional behavioral interventions will be sought to facilitate implementation. For example, in the intervention group, signs in the patient rooms will alert clinical staff to the ongoing experiment and CO_2_ targets.

Following the two-week sham/intervention phase, an anonymous online survey will be sent to ward staff to collect quantitative feedback regarding feasibility and preference to use CO_2_ monitors (using a 10-point Likert scale). The survey will also collect feedback about whether or not staff took action to increase ventilation, their knowledge about the primary CO_2_ target used during the study and the most important challenges to implement a CO_2_ monitoring strategy. We will specifically inquire about staff concerns for patient harms *e*.*g*. from cold or draft discomfort. Hospital-wide COVID-19 clusters will be recorded during the study, but this trial is not intended nor powered for clinical outcomes.

### 2.4. Statistical analysis plan

Our primary hypothesis (primary endpoint) is that the CO_2_ monitors will record less time/day (in minutes) with elevated CO_2_ levels (> 800 p.p.m.) during the intervention period, compared to the sham period. Intervention *vs*. sham control groups will be compared using unpaired t test or Mann-Whitney U test, depending on whether or not the Gaussian distribution of the data is rejected by the D’Agostino-Pearson (omnibus K2) normality test. Secondary outcomes include the time with CO2 > 1000 p.p.m. or > 1400 p.p.m. (which are the built-in cut-off levels for orange and red warning lights on the Aranet^®^ devices), analyzed using similar statistics. Other outcomes include survey responses (see “Intervention” section above).

It is possible (and allowed) that staff applies behaviors that influence CO_2_ levels also in sham rooms. Therefore, the crossover design is applied, with carry-over effects investigated by comparing the Intervention periods to the baseline period. The four periods (baseline, sham, intervention and post-intervention) will be analyzed by one-way ANOVA (or repeated measures ANOVA, if the room-level values are significantly paired) with Dunn’s multiple comparisons post-test (or non-parametric Kruskal-Wallis test if not normally distributed) comparing the intervention period against the baseline, sham and post-intervention periods (3 comparisons). The only exclusion criterion is incomplete occupancy *i*.*e*. when the double-bed rooms are not fully occupied before 12 *a*.*m*., measurements from that day will be excluded. Missing data will not be imputated.

*A priori*, we assume to interpret the results as follows:

- When the intervention *vs*. sham comparison is significant (regardless of the other comparisons), an effect beyond sham will be assumed
  - If this is the case, a significant difference between baseline and intervention will be considered further supportive observational evidence of an intervention carry-over effect (though not required to confirm the randomized intervention *vs*. sham comparison)
  - If this is the case, a significant difference between the intervention and post-intervention phase will be considered supportive observational evidence of a wear-off (or time trend) effect
- If the intervention *vs*. baseline AND intervention *vs*. post-intervention comparison are significant (but not intervention *vs*. sham, which could be masked by carry-over effects), a pre-post effect of the intervention will be assumed.
- If only the intervention *vs*. baseline comparison yields significant results, a hypothesis-generating effect of the intervention could be considered.

Statistical analyses will be performed using GraphPad Prism software. Two-tailed α below 0.05 will be considered as significant. Due to lack of sufficient pilot data for robust power calculation, we assume a conventional moderate effect size (f = 0.25). With an α error probability of 0.05, power of 0.95 and four groups, we calculate total sample size at n=280. With four groups, six sensors, two wards, seven days of measurements and expecting 15 % excluded values due to unoccupied rooms (N = 4 x 6 x 2 x 7 x 0.85 = 285.6), our trial should be powered to detect a moderate effect size. Power calculation was performed using G*Power software version 3.1.9.7 (Kiel University, Germany).

### 2.5. Ethics and trial registration

The study was ethically reviewed by our Institutional Review Board on February 9th, 2021. The Ethical Committee decided that the study did not require informed consent since the design did not qualify as a human clinical trial according to applicable national and European regulations. Only basic demographic data about the room occupants (*i*.*e*. age and sex) as well as bed occupancy will be collected anonymously via the electronic health record database. Still, the head nurses of each ward provided verbal consent to participate voluntarily in the study, and all staff members were free to adhere to recommendations to apply mitigation strategies to avoid high indoor CO_2_ levels or not. There is no funding involved in this trial.

The trial protocol was registered with ClinicalTrials.gov on February 21^st^, 2021 (and published on February 25^th^, 2021) at: https://clinicaltrials.gov/ct2/show/NCT04770597. Previous and future protocol amendments will be registered before publication of the results. The full set of data supporting the trial will be made available to established investigators upon simple request. The authors plan to disseminate the results through international peer-reviewed scientific journals.

## 3. DISCUSSION

We report here the background, rationale and pilot data that led us to the design of the Monitors to Improve Indoor Carbon Dioxide Concentrations in the Hospital (MICH) trial.

There is little doubt that CO_2_ monitors can cheaply and accurately measure CO_2_ levels in a hospital environment. However, like any technology, implementation requires a behavioral change on the part of caregivers. Thus, our trial classifies as “implementation science”, aiming mainly to determine whether nurses and other staff will look at the monitors and increase ventilation, *e*.*g*. by operating available HVAC systems, opening windows and/or altering their daily caregiving routines. Indeed, excessive CO_2_ levels are caused both by raised production by occupants (too many staff members crowding patient rooms for prolonged times) as well as insufficient CO_2_ removal by ventilation. Significant difficulties are expected to implement such behavioral changes, *e*.*g*. because caregivers have many other responsibilities, may resist alterations in their work routines, out of concern that increased ventilation leads to draft or temperature discomfort to patients and staff, or concern that windows must remain closed to prevent patients from falling out.

A criterion of 10 m² space per person is often recommended to allow social distancing in schools, offices and shops. Given the size of our patient rooms and doctor’s office, it is clear that these limits are adequate on average, but that they can be temporarily exceeded by staff occupancy. For example, our doctor’s office size is appropriate for two staff geriatricians, but it does not take into account the 3-6 trainees which also need office space. Hospital space is perennially constrained, and the COVID-19 pandemic has spurred further chaotic reorganizations of wards and offices. Thus, continuous attention to office space occupancy in hospitals in required. Moreover, given secular trends in outdoor CO_2_ concentrations, ideally CO_2_ levels would be monitored simultaneously indoors and outdoors, and referred to in guidelines as p.p.m. above outdoor values, rather than as absolute indoor levels.

A major limitation is that our trial is not designed nor powered to determine whether CO_2_ monitors reduce the risk of hospital-acquired infections including COVID-19. A first step we want to investigate is whether staff can demonstrate behavioral changes in response to the monitors –if not, then any clinical benefit appears unlikely, and simply recommending monitors with the expectation that they by themselves maintain appropriate CO_2_ levels in a clinical environment would be premature. Moreover, CO_2_ is an imperfect proxy of respiratory infection risk. Masks or HEPA filters, loud vocalization [33], [34] or physical distancing for example, affect pathogen transmission risk without influencing CO_2_ levels, while respiratory quotient influences CO_2_ production independent of exhaled air volume. Another limitation of our trial protocol is the single-center open-label design. The latter could not be avoid but makes our findings susceptible to observation bias (Hawthorne effect). Due to lack of sufficient pilot data, a generic power calculation strategy was applied. Still, to the best of our knowledge, this is the first randomized trial aimed to provide a much needed evidence base regarding the effectiveness of CO_2_ monitors in the hospital environment.

## Data Availability

The full set of data supporting the trial will be made available to established investigators upon simple request.

## Acknowledgement

We thank the staff of the geriatrics department for their participation in the study.

